# BIBLIOMETRIC ANALYSIS OF COMPLEMENTARY THERAPY TRENDS FOR MENSTRUATION PAIN

**DOI:** 10.1101/2023.12.27.23300591

**Authors:** Pawestri, Moses Glorino Rumambo Pandin, Esti Yunitasari

## Abstract

**Background:** Research interest in the topic of complementary interventions to treat menstrual pain is increasing. In future research, researchers need information about trends and new things on complementary interventions to treat menstrual pain.

**Purpose:** The research aims to determine trends in the number of publications, journals with the highest number of publications, which fields receive the most publication permits, network visualization, overlay visualization, and density visualization on complementary topics related to intervention dealing with menstrual pain.

**Methods:** A search for publications on trends in complementary interventions to treat primary menstrual pain in adolescents resulted in 23,935 articles, four grants, 2,427 patents, and 42 clinical trials. After filtering through specified criteria, the number of articles obtained was 3,214, 249 patents, and 19 clinical trials.

**Results:** A search for publications on trends in complementary interventions to treat primary menstrual pain in adolescents resulted in 23,935 articles, four grants, 2,427 patents, and 42 clinical trials. After filtering through criteria such as publication year, research categories, publication type, published between 2018-2023, focus in the field of Health sciences, nursing, public health, clinical sciences, and article publication type were included as inclusion criteria of this study. The study obtained 3,214 articles, 249 patents, and 19 clinical trials. The peak of publications regarding trends in complementary interventions to treat primary menstrual pain in adolescents occurred in 2018. Meanwhile, the lowest number was in 2023. Research on complementary intervention trends to overcome primary menstrual pain in adolescents is in medicine, public health, nursing, and other fields such as trade management, tourism and services, human society, law, and legal studies. In addition, there is currently a trend for complementary therapy interventions to treat menstrual pain. Complementary therapies that are still rarely researched are progressive muscle relaxation therapy or herbal ginger to ease menstrual pain.

Apart from that, progressive muscle relaxation therapy can also be used to treat anxiety in teenagers who experience menstrual pain, or the ginger herb has rarely been researched to treat menstrual pain.

**Conclusion:** Trends regarding complementary therapy interventions to treat menstrual pain need a review to find relevant alternative interventions for adolescent development and current digital developments. It is suggested that the next researcher choose a low visualization category theme to look for novelties in further research, one of which is ginger herbal therapy to overcome menstrual pain or interventions to overcome adolescent anxiety when experiencing menstrual pain.

## Introduction

Menstrual pain is a chronic and recurrent pain with the characteristics of lower abdominal pain and pelvic pain or low back pain. The pain can disrupt teenagers’ activities and reduce productivity (Wong et al., 2016). Menstrual pain and symptoms cause school absenteeism in 14–51% of adolescents and disruption of life activities in 15–59% (Parker et al., 2010). Interventions to deal with menstrual pain are increasingly developing, which are adapted to the characteristics of adolescents (Kosanke, 2019), (Pawestri et al., 2023), (Natalia et al., 2020).

There are many complementary therapies to treat menstrual pain, including yoga, acupressure, aroma therapy, relaxation therapy, endorphin massage, aerobic exercise, high-frequency electrical nerve therapy (TENS), or medication (Johnson, 2006), (Ramadhani, 2019), (Tennfjord et al., 2021), (Sari & Usman, 2021), (M.D., 2020), (Zaen, 2021), (Pawestri et al., 2023).

Research interest in the topic of complementary interventions to treat menstrual pain is increasing. Research interest in the topic of complementary interventions to treat menstrual pain is rising, and this happens because complementary therapy is a low-cost and low-cost therapy that has no side effects (Fernández-Martínez et al., 2022).

In future research, researchers need information about trends and new things on complementary interventions to treat menstrual pain.

Trends in complementary interventions to treat menstrual pain can be observed by searching data on Google Trends by typing in the keywords complementary interventions to treat menstrual pain. The search runs from January 2018 to May 2023 via a web search based on all categories. Data was taken on May 29, 2023.

The results of bibliometric analyses may guide future studies by determining the quality and main research areas of existing publications in specific fields. Moreover, bibliometric analysis enables researchers to quickly obtain information from numerous, increasingly published articles about subjects of interest. There is no bibliometric analysis on the publication of research topics on complementary therapy trends for menstrual pain to find trends and novelties related to those topics. This study was conducted to answer questions about:

Q1: How much is the number of publications on complementary intervention to treat menstrual pain?

Q2: How many citations exist on complementary interventions to treat menstrual pain?

Q3: What is the List of journals with the most publications on the topic of complementary intervention to treat menstrual pain

Q4: Which field has the most consent to research the topic of precision medicine and precision health?

Q3: How about network visualization on complementary intervention to treat menstrual pain?

Q4: How do we overlay visualization on the topic of complementary intervention to treat menstrual pain?

Q5: How about density visualization on complementary intervention to treat menstrual pain?

Bibliometric analysis is a method currently trending to study changes in research development, where research topics and researchers are interrelated regarding the research scientific discipline (Naveen Donthu, 2021). Scientific and quantitative methods in assessing a publication can use bibliometric analysis for researchers to see and discover future research development trends (Soytas, 2021). Bibliometric analysis for researchers can help identify emerging themes and future directions of research domains with the help of visualization tools (Lam et al., 2022). Several authors use bibliometric analysis to evaluate information theories in international databases (Lam et al., 2022). Bibliometric analysis intends to obtain descriptive data and findings regarding complementary interventions to treat menstrual pain.

### Purpose

This research aims to determine trends in the number of publications, which fields received the most publication permits, network visualization, overlay visualization, and density visualization on complementary interventions to treat menstrual pain through bibliometric analysis.

## Methods

### Design

There are five study metrics for data analysis: Scientometrics, Bibliometrics, Cybermetrics, Informetrics, and Altmetrics (Chellappandi Ph.D. Assistant Professor & Vijayakumar, 2018). Bibliometrics analysis is more suitable for quantitatively analyzing the distribution of research papers, terms, and keywords in determining research trends (Murugesu et al., 2022). In addition, bibliometric analysis is a research method used in library and information science to evaluate research performance (Syros et al., 2022). Bibliometric analysis is essential in assessing research impact, where studies are ranked based on the citations received (Pahwa et al., 2022).

### Data sources

The data sources used in this study are based on online searches via https://app.dimensions.ai/. Data was collected on May 29, 2023. The literature search used the stages following the Preferred Reporting Items for Systematic Reviews and Meta-Analyses (PRISMA) flowchart (Page et al., 2021).

### Inclusion criteria of articles

The paper was restricted to publications from 2018-2023, focusing on health sciences, nursing, public health, clinical sciences, and publication type. The articles were included as inclusion criteria of this study. After filtering through the requirements of the year of publication, research category, and publication type, with the keywords intervention AND complementary AND pain AND menstruation.

### Selecting data

The stages in PRISMA include identification, filtering, and including, as shown in Figure 1. Stage 1 (Identification) detected 23,935 articles, four grants, 2,427 patents, and 42 clinical trials. After filtering through the criteria, the study obtained 3,214 articles, 249 patents, and 19 clinical trials. 3,214, 249 patents clinical trial records from dimension.ai by considering each of the leading search terms (Complementary interventions to treat menstrual pain), “articles and process document types,” and “all data published in the data range from 2018 to 2023. In stage 2 (filtering), the option “article title, abstract” is selected in the field of each search term, so that • Paper published before years 2018 (n= 861), Not related with the field of Health sciences, nursing, public health, clinical sciences (n=5,800), Not publication in the type of articles (n= 15,680). In phase 3 (inclusive), the final sample yielded 3214 articles. The detailed process is shown in **Figure 1**

**Figure 1.**
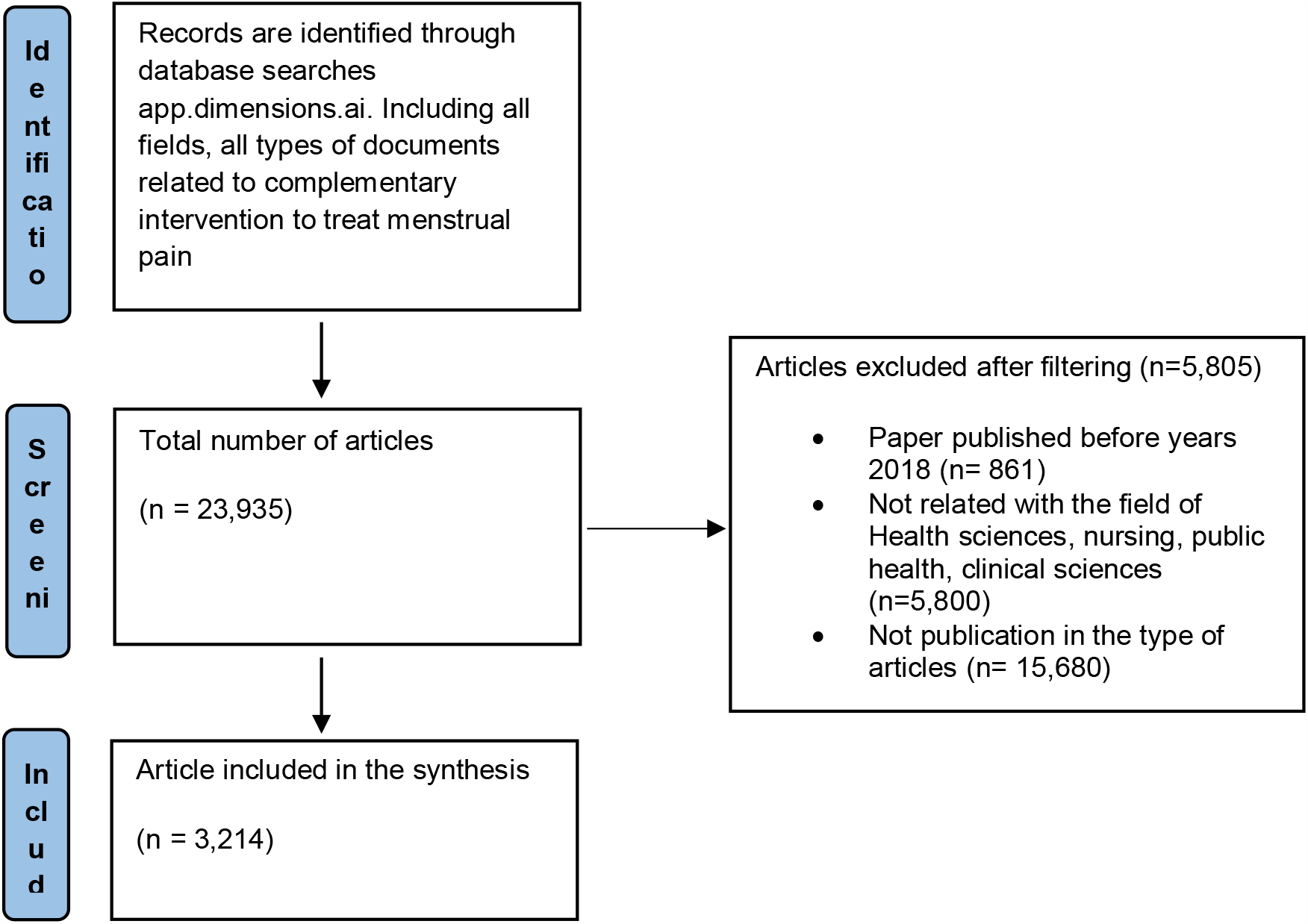
PRISMA Flowchart

### Data analysis

Data were analyzed using VOSviewer. VOSviewer is a computer program for creating and viewing bibliometric maps (van Eck & Waltman, 2010). The type of analysis is selected to create a map based on text data. In this study, the investigation was reviewed by co-occurrence and co-authors.

### Co-occurrence procedures

Co-occurrence analysis uses several procedures: selecting the data source and reading data from the reference management file. Determines multiple terms from the specified fields, then extracts them from the title and abstract fields. The minimum number of occurrences is 15 out of 175. the threshold is the minimum number of term appearances (Suparman, 2023).

### Co-authors procedures

The co-author analysis procedure went through several stages. First, choose the type of data, namely, create a map based on bibliographic data. Second, select the option to create a co-authorship map based on bibliographic data. Third, choose the data source: reading data from the reference manager file. The supported file type is ris. Fourth, select the analysis and calculation method; the kind of analysis is co-authorship, and the calculation method is complete calculation. Select a threshold: the maximum number of documents for an author is 3. Of the 3,214 authors, 34 met the point. Select authors: For each of the 34 authors, the total strength of co-authoring relationships with other authors is calculated. The author with the most significant total link power will be selected. The number of selected authors was 34.

## Results

### Publication searches related to trends in complementary intervention to treat menstrual pain in adolescents

**Figure 2.**
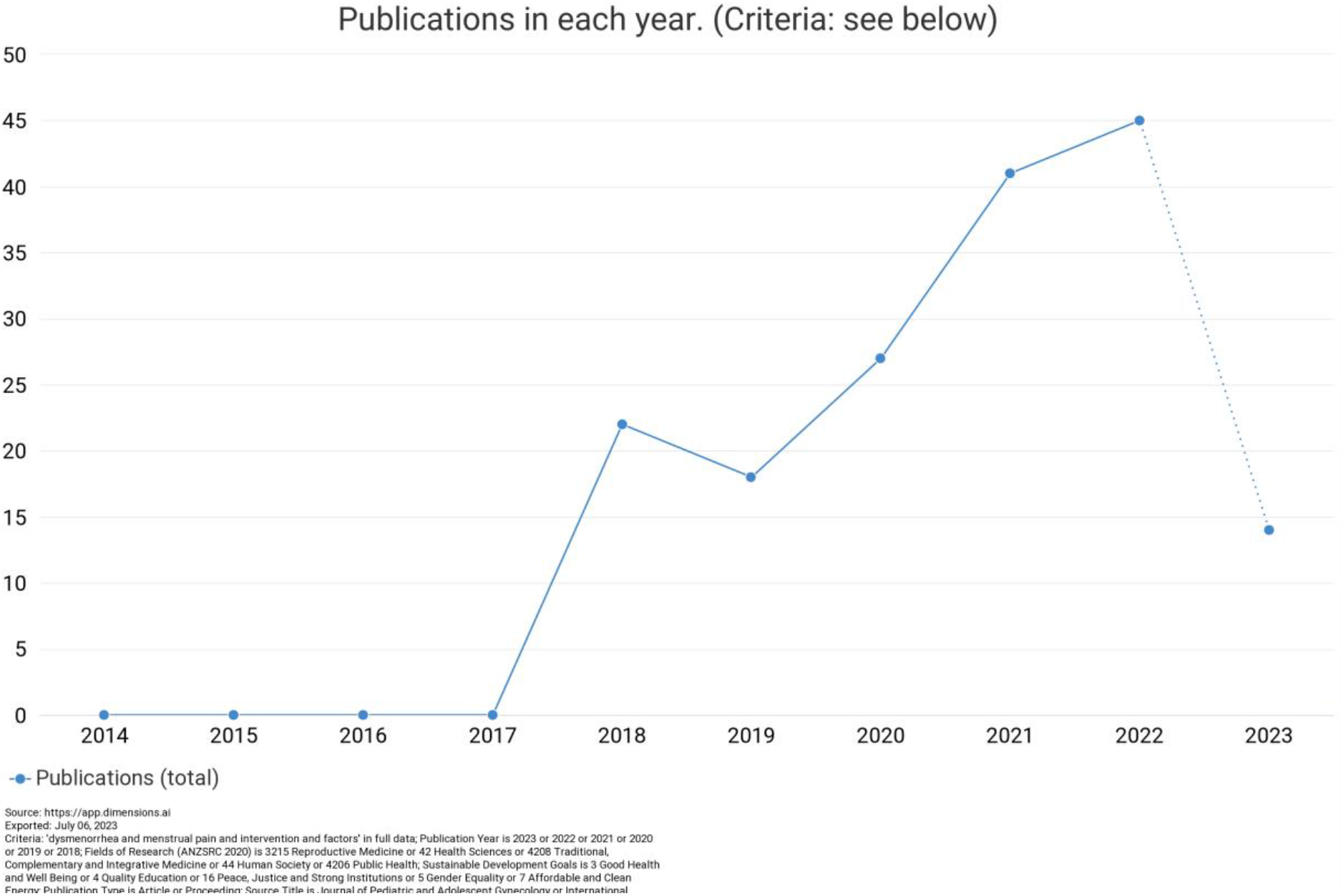
Number of publications on complementary intervention to treat menstrual pain from 2018-2023 (sources: https://app.dimensions.ai/)

### Publication searches related to trends in complementary intervention to treat menstrual pain in adolescents by Research Category

**Figure 3.**
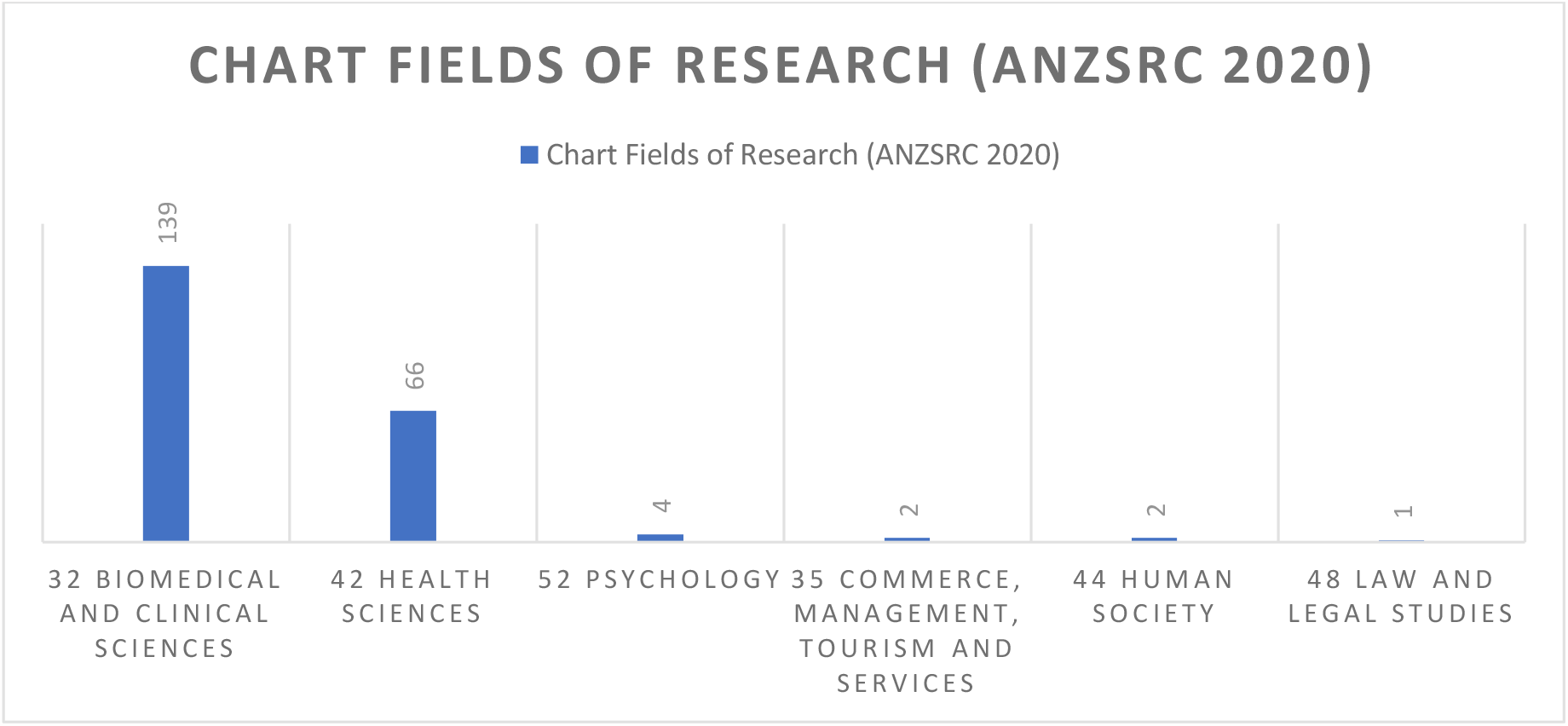
Number of publications of each research category (sources: https://app.dimensions.ai/)

### Publication searches related to the List of journals with the most publications on complementary interventions to treat menstrual pain. Network visualization of precision health and precision medicine concepts in publications

**Figure 4.**
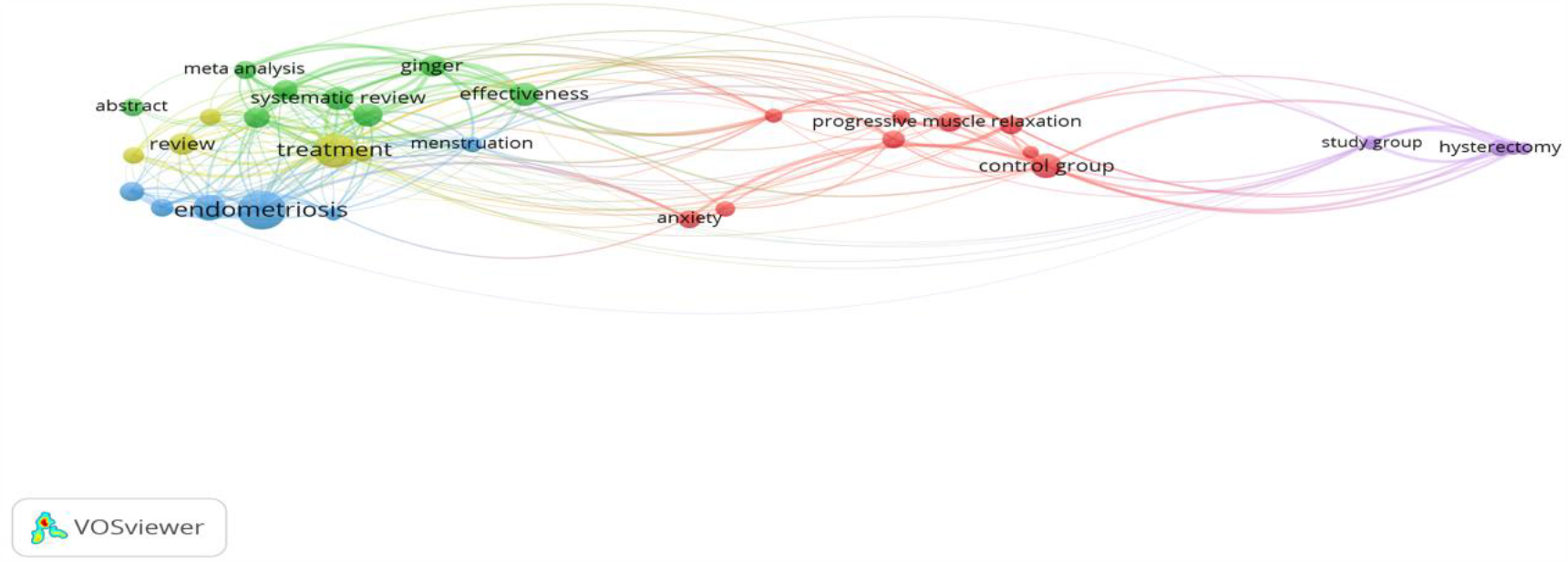
Network visualization (sources: VOSviewer)

### Publication searches related to Overlay visualization of complementary intervention to treat menstrual pain in publications

**Figure 5.**
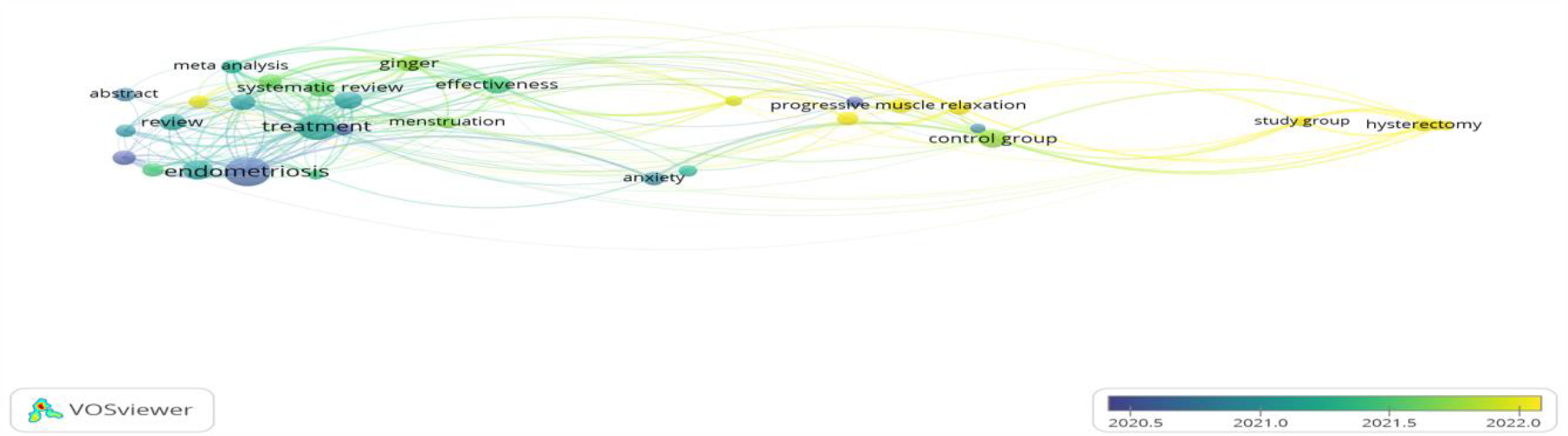
Overlay visualization (sources: VOSviewer)

### Publication searches related to Density visualization of complementary intervention to treat menstrual pain in publications

**Figure 6.**
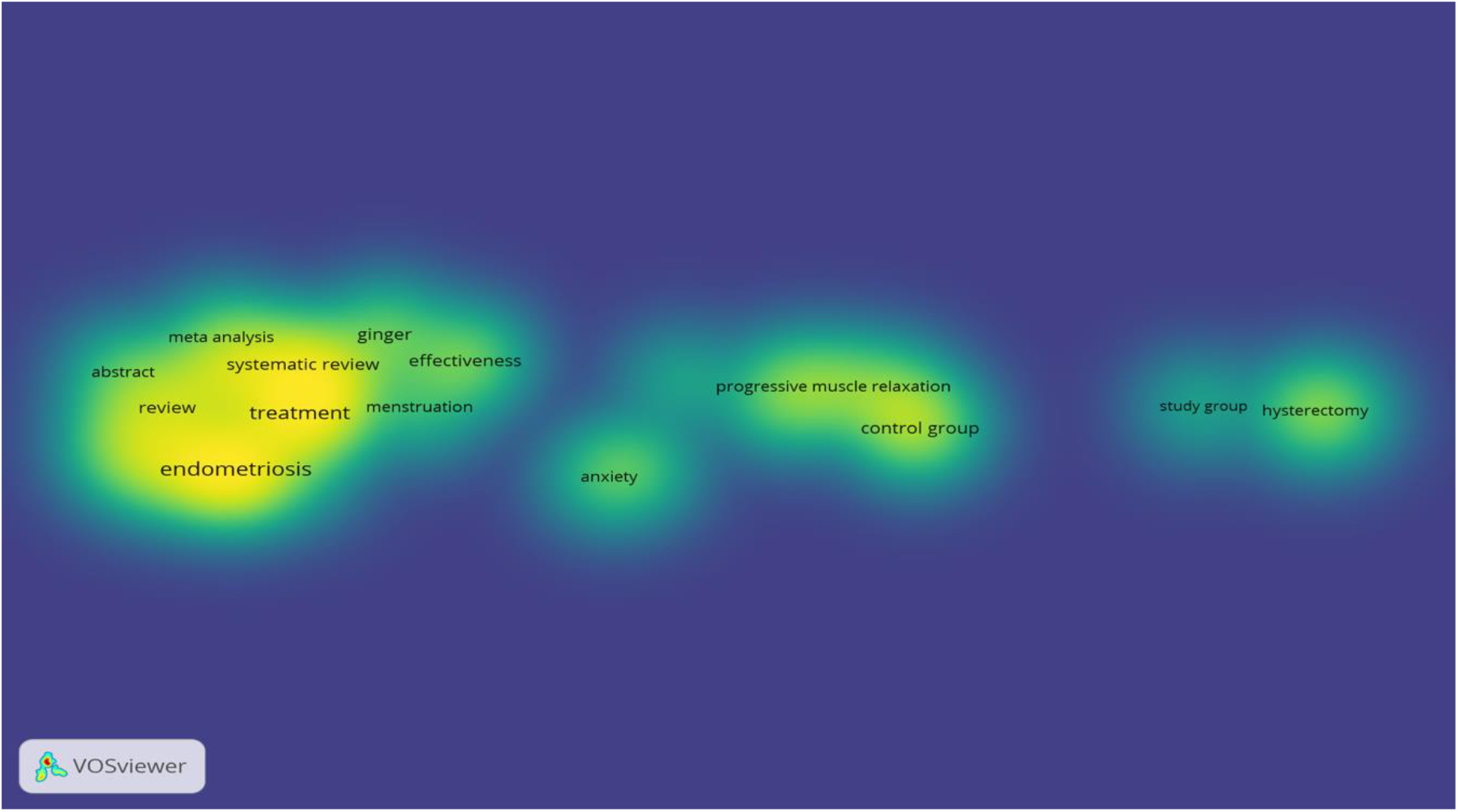
Density visualization (Sources: VOSviewer)

## Discussion

Searching for publications related to trends in the number of publications, trends in the number of citations, journals with the highest number of publications, which fields received the most publication permits, network visualization, overlay visualization, and density visualization on the topic of complementary interventions for the treatment of menstrual pain resulted in the publication of 23,935 articles, four grants, 2,427 patents, and 42 clinical trials. After filtering through specified criteria, the number of articles obtained was 3,214, 249 patents, and 19 clinical trials.

The peak of publications regarding complementary interventions to treat menstrual pain occurred in 2022 and decreased in 2019, while the lowest publications occurred in 2023 because it was still in the continuing period. After filtering through specified criteria, the number of articles obtained was 4060 from 2017-2023. The highest number of publications regarding complementary interventions to treat menstrual pain increases every year. In 2022, there will be 2,965 articles, while the lowest number will be in 2023, with 530 articles. During the 2019 coronavirus pandemic, there was a significant decrease in publications on complementary interventions to treat menstrual pain. The number is related to the pandemic at the end of 2019-2021 (Lambovska et al., 2021). Researchers from the beginning of 2020 to the end of 2021 focused more on research related to COVID-19 (Surmiak et al., 2022).

On the other hand, from mid-2022 to the end of 2022, there was a significant increase in research publications on the topic of complementary interventions to treat menstrual pain, and this will still increase in 2023 (Lee et al., 2022; Mitchell et al., 2022). The publication is closely related to the decline in COVID-19 cases at the end of 2022, so researchers focus the study on other topics to reduce direct contact with respondents. There will still not be many publications in 2023 regarding this topic because the literature search related to this topic was finished in May 2023 and didn’t reach the end of 2023, so many publications are still related to complementary interventions to overcome menstruation pain. Publication trends from year to year are presented in Figure 2.

Research on complementary interventions to treat menstrual pain is not only carried out by the fields of medicine, psychology, public health, or nursing but also by the fields of trade management, tourism and services, human society, and legal science. The health sector mostly dominates the number of publications related to complementary interventions to treat menstrual pain. Still, other fields, such as trade management, tourism and services, public health, and legal studies, also explore the topic of complementary interventions to deal with menstrual pain (Armour et al., 2019).

There are 33 biomedical and clinical sciences journals with 139 articles, 42 health sciences journals with 66 titles, 52 psychology journals with four titles, 35 commerce management, tourism, and services journals with two articles, 44 human society journals with two articles, and 48 law and legal studies contains 1 article. It’s in Figure 3.

The Management sector discusses menstrual pain because when women experience menstrual pain, productivity will be disrupted, one of which is employees taking leave when experiencing menstrual pain. Implementing sick leave provisions requires a doctor’s letter to prove that the female worker is menstruating (Law of the Republic of Indonesia No. 13 of 2003 concerning Employment, 2003:19).

The tourism and services sector is also interested in treating menstrual pain because this profession will experience problems when its customers experience menstrual pain, which results in the customer’s journey becoming uncomfortable. After all, this requires intervention that can be done to overcome it. One of the factors that influence menstrual pain is that excessive activity can cause menstrual pain (Rostami et al., 2006). Women typically refrain from traveling during menstrual pain, which has an impact on the tourism sector.

From the analysis results in Figure 4, exciting things were found, including one of the interventions to overcome menstrual pain with ginger. There is also progressive muscle relaxation, which can also be used to overcome menstrual pain and anxiety when experiencing menstrual pain. Many make it systematic—review and meta-analysis regarding implementation to overcome menstrual pain. One complementary intervention to deal with menstrual pain that is developing is using ginger. Ginger alternative therapy can reduce dependence on synthetic drugs to control Dysmenorrhea (Gurung et al., 2022). Complementary intervention research currently being carried out uses progressive muscle relaxation to treat menstrual pain. Progressive muscle relaxation affects menstrual pain (Pawestri et al., 2023).

The research that is starting to be developed is progressive muscle relaxation to overcome anxiety in women who experience menstrual pain. Regular progressive muscle relaxation has been shown to reduce anxiety, depression, and stress symptoms (Jacob & Sharma, 2018).

Research that is currently of interest regarding complementary interventions to reduce menstrual pain is meta-analysis and systematic reviews. The benefits of this therapeutic modality for treating menstrual pain and the low risk of reported side effects are that women consider using complementary interventions that can be done alone and do not have side effects (Jo & Lee, 2018),(Abaraogu et al., 2016).

From the results of the analysis in Figure 5 Overlay visualization, it was found that progressive muscle relaxation is a trending intervention to treat dysmenorrhea pain which is of interest to researchers because the results of the analysis show that progressive muscle relaxation interventions are usually used in hospitals to treat pain or anxiety (Rahman et al., 2022),(Ismail & Elgzar, 2018),(Purnomo et al., 2021),(Wirakhmi et al., 2020),(Ovgun & Tuzun, 2023),

Progressive muscle relaxation can be used in the community, especially for teenagers who experience menstrual pain. Progressive muscle relaxation is a therapy that is easy to do yourself and has no side effects, so many researchers have developed progressive muscle relaxation. Progressive muscle relaxation intervention can overcome several problems by providing comfort and relaxing the muscles so that oxygen circulation in the endometrium is smooth. That discomfort can be resolved (Çelik & Apay, 2021).

From the results of the analysis in the visualization of Figure 6 Overlay, it is found that there are bright and dark images, which means that the image looks bright, meaning the research is getting more saturated. If the image looks dim, then the research is rarely researched. From Figure 6 overlay, it can be concluded that research on complementary therapies that are rarely studied to treat menstrual pain in adolescents includes progressive muscle relaxation and ginger. Researchers rarely analyze the anxiety problems experienced by teenagers who experience menstrual pain. Complementary yoga relaxation therapy can be used to overcome anxiety in teenagers who experience menstrual pain (Awaliyah & Pragholapati, 2021),(Jadhao, 2019),(Kucukkelepce et al., 2021).

Progressive muscle relaxant therapy or ginger therapy can be used to treat menstrual pain but is rarely used in research to treat anxiety. Progressive muscle relaxant to treat anxiety (Liu et al., 2020). Ginger therapy can be used to treat menstrual pain in teenagers (Mozafari et al., 2018), (Jenabi, 2013), (Gutman et al., 2022). Ginger therapy is a progressive muscle relaxant capable of being used to treat menstrual pain.

## Implication and limitations

Although this research contributes to increasing insight into complementary therapy interventions carried out to treat menstrual pain from 2018-2023 via app.dimension.ai, this research may have limitations. The app.dimensional.ai database is still updating new publications gradually. Therefore, bibliometric analysis of complementary therapy interventions carried out to treat menstrual pain still needs to be reviewed in the next few years. Further research is needed to add a broader database for a comprehensive understanding of complementary therapy interventions to treat menstrual pain.

## Conclusion

The peak of publications regarding complementary therapy interventions to treat menstrual pain increased every year from 2018 to 2022. The downward trend only occurred in 2019 in the Covid era, and in 2020 there was another increase. Research on non-pharmacological therapeutic interventions to treat menstrual pain is not only carried out by the fields of medicine, public health, or nursing but also by other fields. Fields include psychology, commerce, management, tourism and services, human society, law, and legal studies. The field is closely related to using data technology such as artificial intelligence, big data, and genetic analysis. Apart from that, the health trend in dealing with menstrual pain is not only pharmacological therapy but also non-pharmacological therapy, in this case, complementary therapy. Complementary progressive muscle relaxation therapy and ginger therapy need to be developed to treat menstrual pain. The anxiety problem of teenagers who experience menstrual pain requires intervention, one of which is progressive muscle relaxation.

## Data Availability

All data produced in the present study are available upon reasonable request to the authors

https://drive.google.com/file/d/1Hb0mtiofTAGAEkPH9QWInYWQemsOzcTH/view?usp=sharing

